# The cost-effectiveness of small-quantity lipid-based nutrient supplements for prevention of child death and malnutrition and promotion of healthy development: modeling results for Uganda

**DOI:** 10.1101/2022.05.27.22275713

**Authors:** Katherine P Adams, Stephen A Vosti, Charles D Arnold, Reina Engle-Stone, Elizabeth L Prado, Christine P Stewart, K Ryan Wessells, Kathryn G Dewey

**Author notes:** Corresponding author Institute for Global Nutrition, University of California, Davis, One Shields Avenue, Davis, CA 95616. **Authorship** KGD conceptualized the study. All authors designed the study. KPA and SAV developed the cost model. KPA and CDA developed the effectiveness model. KPA and KGD wrote the first draft of the manuscript. All authors contributed to data interpretation and revisions of the manuscript, and read and approved the final manuscript. **Ethical Standards Disclosure** Not applicable.

## Abstract

**Objective:** Recent meta-analyses demonstrate that small-quantity lipid-based nutrient supplements (SQ-LNS) for young children significantly reduce child mortality, stunting, wasting, anemia and adverse developmental outcomes. Cost considerations should inform policy decisions. We developed a modeling framework to estimate the cost and cost-effectiveness of SQ-LNS and apply the framework in the context of rural Uganda.

**Design:** We adapted costs from a costing study of micronutrient powder (MNP) in Uganda, and based effectiveness estimates on recent meta-analyses and Uganda-specific estimates of baseline mortality and the prevalence of stunting, wasting, anemia, and developmental disability.

**Setting:** Rural Uganda.

**Participants:** Not applicable.

**Results:** Providing SQ-LNS daily to all children in rural Uganda (>1 million) for 12 months (from 6-18 months of age) via the existing Village Health Team system would cost ∼$52 per child (2020 US dollars), or ∼$58.7 million annually. Annually, SQ-LNS could avert an average of >242,000 disability adjusted life years (DALYs) as a result of preventing 3,689 deaths, >160,000 cases of moderate or severe anemia, and ∼6,000 cases of developmental disability. The estimated cost per DALY averted is $242, which is considered “very cost effective” relative to the Uganda per capita GDP of $822.

**Conclusions:** In this context, SQ-LNS may be more cost-effective than other options such as MNP or the provision of complementary food, although the total cost for a program including all age-eligible children would be high. Strategies to reduce costs, such as targeting to the most vulnerable populations and the elimination of taxes on SQ-LNS, may enhance financial feasibility.

## Introduction

Small-quantity lipid-based nutrient supplements (SQ-LNS) were developed to prevent malnutrition among vulnerable populations. These fortified food-based products (typically including vegetable oil, peanut paste, and milk powder) deliver vitamins and minerals as well as essential fatty acids and small amounts of energy (∼100-120 kcal/day) and protein (1). Since the development of SQ-LNS ∼15 years ago, a strong evidence base has been established demonstrating that SQ-LNS for children 6-24 months of age reduces child mortality, stunting, wasting, and anemia and promotes healthy development in efficacy trials as well as effectiveness trials in programmatic settings (2-6). As a result, SQ-LNS was listed in the 2021 Lancet Series on Maternal and Child Undernutrition as an intervention with a strong evidence base supporting implementation (7).

Alongside robust evidence of effectiveness, informed policy decisions made by governments, aid organizations, and other agencies regarding their portfolios of nutrition interventions also require information about the cost and cost-effectiveness of various options. There is very limited evidence on the cost, and no evidence on cost-effectiveness, of providing SQ-LNS to young children; this study is a first step toward filling that gap. Our goal was to develop a modeling framework to estimate the potential effects, costs, and cost-effectiveness of SQ-LNS. We selected rural Uganda for this first modeling exercise because detailed data were available on the costs of delivering a similar product, micronutrient powder (MNP), to young children in that setting (8). Our specific objectives were to 1) develop a cost model, adapted from the MNP study, to estimate the annual total cost and cost per child of providing SQ-LNS, 2) develop an effectiveness model to estimate the effects of SQ-LNS on child mortality, anemia, developmental disability, stunting, and wasting and 3) translate those effects into cost per disability-adjusted life years (DALYs) averted, per death averted, and per case of anemia, developmental disability, stunting, and wasting averted in rural Uganda.

## Methods

We developed models to estimate the cost, cost-efficiency (cost per child), and cost-effectiveness of SQ-LNS for young children. The modeling was based on providing 12 months of daily SQ-LNS (20 g/day), beginning at 6 months of age and ending at 18 months of age, to all young children residing in rural districts of Uganda. The models were based on SQ-LNS being delivered via the existing Village Health Team (VHT) community health worker program. The VHTs in Uganda rely on volunteer health workers to deliver basic health and nutrition products and services, provide education, and make referrals (9). Our modeling assumed that current VHTs could, with additional monetary incentives (described below), take on the delivery of SQ-LNS to households in addition to their current activities. We modeled costs and effects over the period 2021-2031, assuming one year of start-up and 10 years of SQ-LNS provision.

### Costs

The cost model was developed based on an actual costing study conducted in one rural district in Uganda (Namubumba District) to estimate the cost of providing MNP to children 6-24 months of age (8). The MNP costing study was designed to compare the cost and cost-efficiency of delivering MNP via community-versus facility-based platforms. The study found that the community-based platform (that is, via VHTs) was more cost-efficient (lower cost per child reached), so we adopted the community-based platform for our SQ-LNS cost model.

We estimated economic costs from a societal perspective, meaning all costs were accounted for, regardless of who incurred them and including both the costs needed to finance the program as well as opportunity costs (e.g., the opportunity cost of caregivers’ time to participate in the intervention and volunteer health worker time). Following the activity-based approach used in Schott, Richardson (8), we estimated the cost of one year of start-up activities (primarily social and behavior change communication (SBCC) and capacity building) and recurring activities over a 10-year time horizon, which included caregiver opportunity costs, SBCC, logistics, capacity building, operational monitoring and evaluation (M&E) costs, overhead and capital, VHT incentives, SQ-LNS product costs, SQ-LNS international shipping and handling and customs clearance, and domestic SQ-LNS transport, storage, and handling.

The product cost of SQ-LNS was based on the price of a carton (546 20-gram sachets) of SQ-LNS published in the UNICEF Supply Catalog (https://supply.unicef.org/s0000245.html) in March of 2022 and adjusted to 2020 US dollars. Although local production is feasible in several African countries, we assumed international production because cost information was more reliable. The cost of SQ-LNS, which was applied to each of the 10 years of intervention, was $33.30/carton (2020 US dollars), or $3.06/kg, which is ∼$0.061 per 20-gram sachet. International shipping and handling costs were estimated at US$0.31/kg based on the default estimate in the FACET tool for shipping and handling of SQ-LNS on the South East Africa trade route (http://facet4snf.org/). Customs clearance costs were estimated at 18% for a value added tax + 33% withholdings tax (10), or ∼ US$1.01/kg. Domestic transport, storage, and handling was estimated to cost US$0.17/kg, based on the default estimate for Uganda in the FACET tool.

Given documented challenges faced by the volunteer VHT system that underlies the community-based distribution platform in rural Uganda, such as inadequate support and high rates of attrition (9, 11), we assumed that the successful addition of SQ-LNS to the products and services delivered by VHTs would require a strengthening of the VHT system. Informed by a Uganda pilot performance-based incentive system (12), we modeled an additional performance-based monetary incentive to VHT health workers of US$0.44 per delivery of SQ-LNS, assuming SQ-LNS was delivered to households every three months.

All other cost estimates were adapted from the MNP costing study (8) by first identifying the base cost of each input (personnel, transport, materials, in-kind incentives) for each activity (plus overhead and capital costs) based on the costs actually incurred in Namutumba. We converted all base costs to 2020 US dollars using the Bureau of Economic Analysis implicit price deflators for gross domestic product (Supplemental Table 1).

We then extrapolated those base costs to all other rural districts in Uganda via a set of extrapolation indices defined relative to Namutumba. The extrapolation indices were: 1) a spatial index based on the area of each district relative to Namutumba, 2) a child population index based on the number of eligible children per district, 3) a VHT population index based on the number of VHTs per district, and 4) a VHT-child population index based on the ratio of VHTs to eligible children per district compared to Namutumba (Supplemental Figure 1). We used these indices to generate district-specific costs for each rural district that reflected differences in each district relative to Namutumba that would likely impact costs. For example, we assumed the cost of transport required for capacity building activities would vary based on the area of each district, so if the area of Namutumba was approximately 820 km^2^ and the area of another rural district was approximately 1,585 km^2^, the index for that district was defined as (1,585/820)=1.93, and the cost for transport associated with capacity building in that district was 1.93 times higher than in Namutumba District.

### Effectiveness

The effectiveness model was designed to estimate annual deaths averted and annual cases of moderate or severe anemia, developmental disability, stunting, and wasting averted attributable to SQ-LNS over a 10-year time horizon (Table 1). The model was also used to estimate DALYs averted, as described below. We applied the effect of SQ-LNS on each of these outcomes to specific age bands depending on the expected timing of effects relative to supplementation. We applied mortality, stunting, and wasting reductions through the full period of supplementation (6-18 months of age). We applied reductions in moderate and severe anemia from 9 to 24 months of age, assuming that changes in anemia status would become evident after ∼3 months of supplementation and would persist for six months post-supplementation. Finally, we modeled the onset in reductions in developmental disability during the 18- to 24-month age range. We used the Lives Saved Tool (LiST) population projections and the UN World Urbanization Prospects to estimate rural child populations in 2022-2031 (Supplemental Table 2).

**Table 1.**
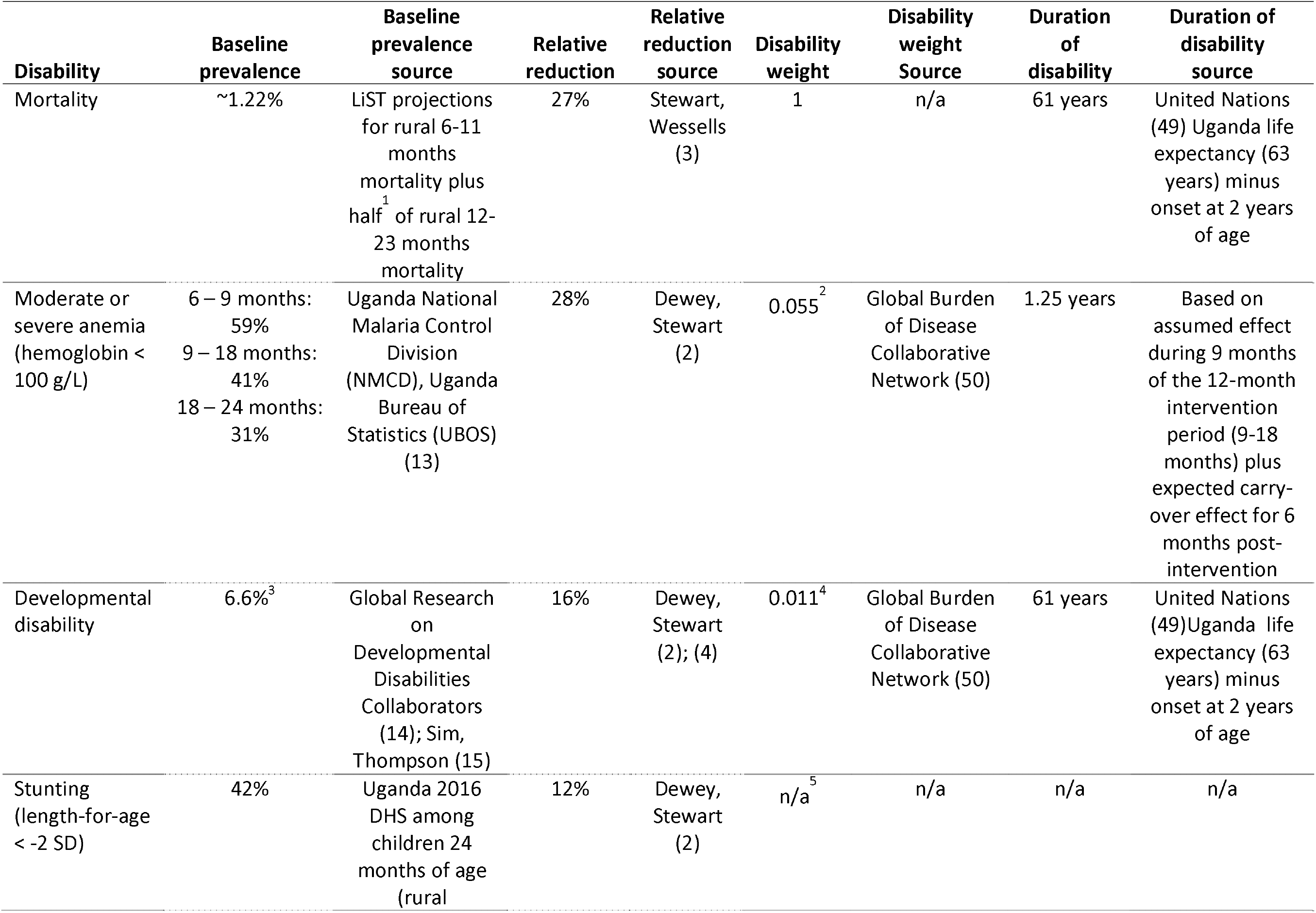
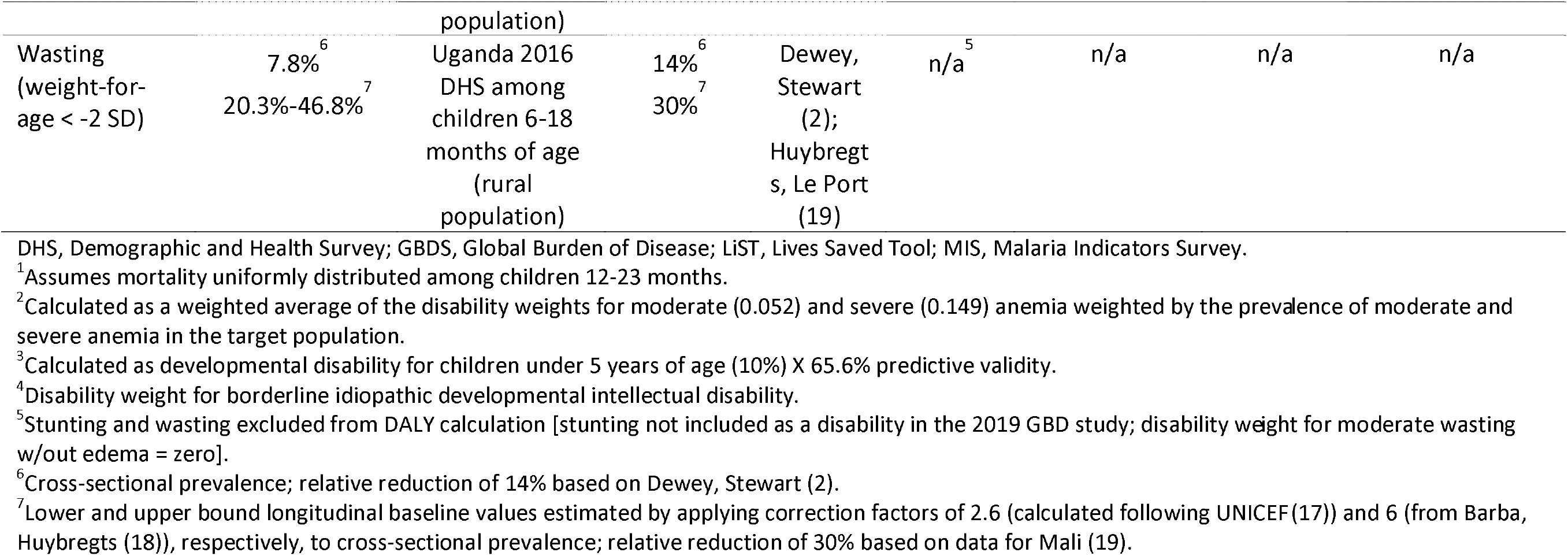
Effectiveness modeling parameter values and data sources

For each outcome, modeled effectiveness of the intervention was calculated as the baseline prevalence of the adverse outcome, multiplied by the relative reduction as a result of SQ-LNS, multiplied by the relevant population size. The baseline mortality rate (∼1.22%) was based on LiST subnational projections of mortality among rural Ugandan children age 6-11 months plus half of projected mortality among rural children 12-23 months (Table 1). We used data from the most recent Malaria Indicator Survey (2018-2019) to estimate the baseline prevalence of moderate and severe anemia among rural children 6-9 months (59%), 9-18 months (41%), and 18-24 months (31%) (13). The baseline prevalence of developmental disability of 6.6% was based on the estimate of 10% prevalence of developmental disability among children under 5 years of age in low- and middle-income countries (LMICs) (14) multiplied by the predictive validity of 65.6% for a score in the lowest decile of the early childhood MacArthur-Bates Communicative Development Inventories (CDI) to predict later language delay (15) (the CDI was the most commonly used tool to assess the effect of SQ-LNS on language development among the studies in the most recent meta-analysis of SQ-LNS) (4). We used data from the 2016 Uganda Demographic and Health Survey (DHS) to estimate the baseline prevalence of stunting of 42% (16). For wasting, we used a cross-sectional prevalence of 7.8% based on data from the 2016 DHS (16). We also estimated a lower and upper bound for the longitudinal prevalence of wasting by applying correction factors of 2.6 (calculated following UNICEF (17)) and 6 (from Barba, Huybregts (18)), respectively, to the cross-sectional baseline prevalence.

We modeled a 27% relative reduction in mortality based on a meta-analysis of randomized controlled trials assessing the impact of SQ-LNS on all-cause mortality in children 6-24 months (3). Relative reductions in moderate-to-severe anemia (28%), developmental delay, i.e., scoring in the lowest decile for language (16%), stunting (12%), and cross-sectional prevalence of wasting at endline (14%) were based on individual participant data meta-analyses (2). We modeled a 30% relative reduction in longitudinal prevalence of wasting based on findings from a cluster randomized controlled trial in Mali (19), assuming that the impacts of SQ-LNS would be similar in Uganda.

We estimated DALYs as the sum of years of life lost (YLL) due to mortality and years lived in disability (YLD) due to moderate or severe anemia and developmental disability. YLL and YLD were calculated as the product of cases of disability, the duration of disability, and the disability weight (Table 1). We did not apply age weighting. DALYs averted were calculated as the difference in DALYs with and without the SQ-LNS intervention. We did not include stunting or wasting in our DALY estimates because stunting is not included as a disability in the 2019 GBD study, and the disability weight for moderate wasting without edema is zero. Mild anemia was not included because estimates of the impact of SQ-LNS on reductions in the prevalence of mild anemia alone were not available from the meta-analyses, and the disability weight for mild anemia was close to zero (0.004).

### Discounting

We calculated costs, effectiveness, and cost-effectiveness under two scenarios. The first assumed a discount rate of zero following the recommendation in Murray, Vos (20) and the second a discount rate of 3% as recommended in World Health Organization (21) and Sanders, Neumann (22). A discount rate greater than zero implies that costs and effects incurred in the future have a lower value than those same costs and effects had they been incurred today. Discounted DALYs averted were calculated following Fox-Rushby and Hanson (23) without age weighting.

### Sensitivity Analyses

We conducted several analyses to evaluate the sensitivity of our results to key assumptions and influential parameter values. First, based on the result of a sensitivity analysis scenario in Stewart, Wessells (3) in which passive control arms were excluded from the analysis, we modeled a scenario in which the relative reduction in mortality was 18% rather than 27%. We also modeled several different cost scenarios that included (1) elimination of the customs clearance costs for SQ-LNS, (2) a 10% decrease in the price of SQ-LNS, (3) a 10% increase in the price of SQ-LNS, (4) lower VHT monetary incentives (reduced from $0.44 to $0.29 per delivery of SQ-LNS), or (5) higher VHT monetary incentives (increased from $0.44 to $0.87 per delivery of SQ-LNS). We also modeled “best” and “worst” case scenarios in which sensitivity analyses parameter values that would improve the cost-effectiveness of SQ-LNS were modeled simultaneously and those that would reduce cost-effectiveness were modeled simultaneously.

### Patient and Public Involvement

Because our study was based on secondary data, there was no patient or public involvement in this research.

## Results

### Costs

The estimated annual average cost of providing SQ-LNS for 12 months to all children starting at age 6 months in rural districts of Uganda via the VHT system was ∼$58.7 million (2020 US dollars), or $52 per child (Table 2; Supplemental Table 3 for costs by year). Approximately $22 of the total cost per child was the cost of the product, ∼$11 was international shipping and handling, customs clearance, and domestic transport, storage, and handling, and ∼$19 was non-product programmatic costs. The cost of the product represented the highest share of the total annual average cost (42.5%), followed by international shipping and handling and customs clearance for SQ-LNS (18.7%), capacity building (15.3%), and logistics (6%) (Figure 1).

**Table 2.** Estimated cost and cost-efficiency of providing daily SQ-LNS to all children in rural Uganda (2020 US Dollars)

| **Cost basis** | **Product cost** | **Product shipping, taxes, and domestic transport cost<sup>1</sup>** | **Non-product programmatic cost** | **Total cost** |
| --- | --- | --- | --- | --- |
| Per sachet | \$0.06 | \$0.03 | \$0.05 | \$0.14 |
| Per child<sup>2</sup> | \$22.32 | \$11.05 | \$19.13 | \$52.49 |
| Average per year (all children)<sup>3,4</sup> | \$24,959,289 | \$12,352,734 | \$21,390,199 | \$58,702,222 |

<sup>1</sup>Product shipping, taxes, and domestic transport includes the cost of international shipping and handling, customs clearance, and domestic transport, storage, and handling.
<sup>2</sup>Assuming daily supplementation for 12 months.
<sup>3</sup>Based on providing SQ-LNS to an average of 1,118,340 children per year.
<sup>4</sup>Average cost per year on the basis of non-product programmatic cost and total cost include annualized start-up costs.

**Figure 1.**
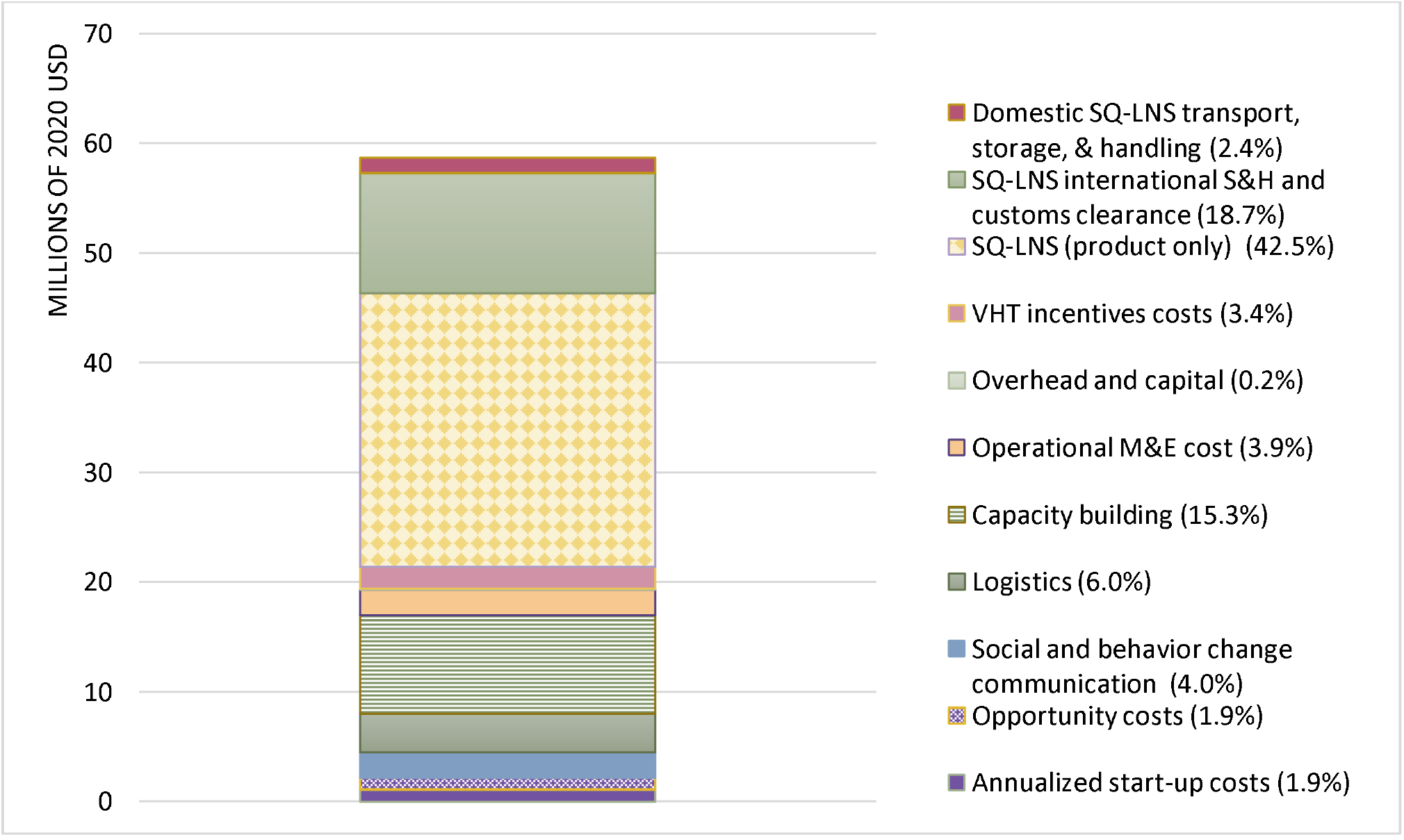
Estimated average annual cost, by activity, of daily provision of SQ-LNS to all children age 6-18 months in rural Uganda delivered via village health teams. Costs in millions of 2020 US dollars. M&E, monitoring and evaluation; SQ-LNS, small-quantity lipid-based nutrient supplements; S&H, shipping and handling; VHT, village health team.

### Effects and cost-effectiveness

We estimated that the provision of SQ-LNS in rural Uganda could avert an annual average of 242,292 DALYs as a result of 3,689 annual child deaths averted, over 160,000 cases of moderate or severe anemia averted, and almost 6,000 cases of developmental disability averted (Table 3). In addition, we estimated that SQ-LNS could annually avert an average of 55,858 cases of stunting, over 12,000 cases of wasting based on cross-sectional estimates, and between 68,040 and 157,015 cases of wasting based on longitudinal estimates.

**Table 3.** Estimated effectiveness and cost-effectiveness of providing daily SQ-LNS to all children in rural Uganda

| **Outcome** | **Annual average effectiveness over 10 years of intervention** | **Undiscounted cost-effectiveness<sup>1,2</sup>** | **Discounted cost-effectiveness<sup>1,3</sup>** |
| --- | --- | --- | --- |
| DALYs averted | 242,202 | \$242 | \$413 |
| Deaths averted | 3,689 | \$15,914 | \$15,538 |
| Cases of anemia averted | 160,267 | \$366 | \$357 |
| Cases of developmental disability averted | 5,905 | \$9,941 | \$9,684 |
| Cases of stunting averted | 55,828 | \$1,051 | \$1,024 |
| Cases of wasting averted (14% RR) | 12,212 | \$4,807 | \$4,683 |
| Cases of wasting averted (30% RR)<sup>4</sup> | 68,040 - 157,015 | \$374-\$863 | \$364-\$840 |

RR, relative reduction.
<sup>1</sup>All estimates of cost-effectiveness presented in 2020 US dollars.
<sup>2</sup>Cost-effectiveness (cost per unit of effectiveness) over modeled intervention period of 2021-2031.
<sup>3</sup>Discounted cost-effectiveness estimates based on discounting both costs and effects at 3%.
<sup>4</sup>Based on lower and upper bound longitudinal prevalence of wasting estimates of 20.3% and 46.8%

We estimated that the undiscounted cost-effectiveness of SQ-LNS over the 10-year intervention time horizon was $242 per DALY averted, or $413 per DALY averted with costs and effects discounted at 3% (Table 3). For individual outcomes, undiscounted cost-effectiveness ranged from $366 per case of moderate-to-severe anemia averted to $15,914 per death averted.

In the sensitivity analyses (Figure 2), the undiscounted cost per DALY averted ranged from $195-$382 under the best- and worst-case scenarios, and the cost per death averted ranged from $12,823-$26,003. The cost per DALY averted and per death averted were most sensitive to changing the assumed mortality reduction from 27% to 18%.

**Figure 2.**
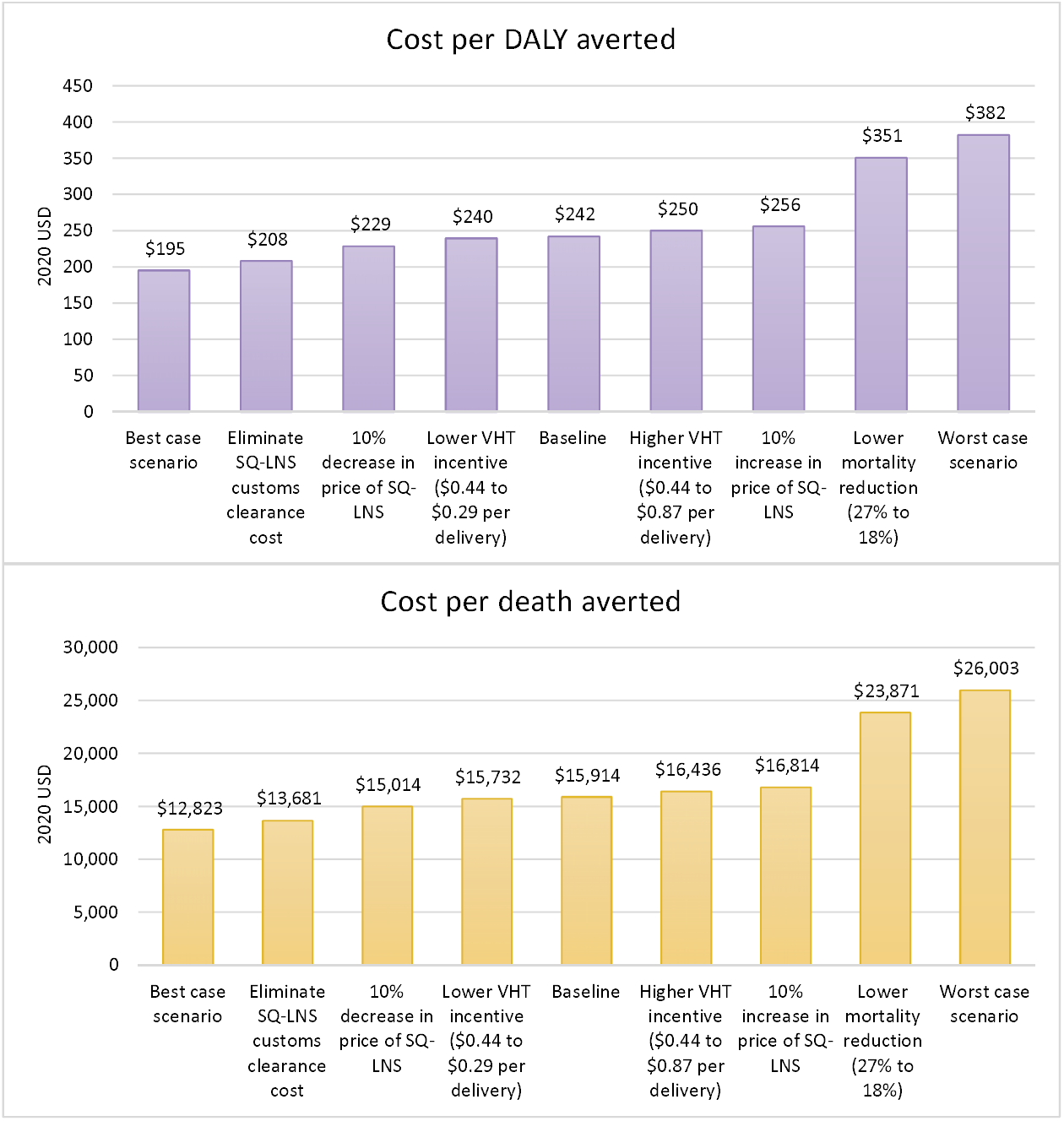
Cost per disability adjusted life year (DALY) averted (top panel) and cost per death averted (bottom panel) under each sensitivity analysis scenario. The best-case scenario simultaneously models the elimination of customs clearance costs, a 10% decrease in price of SQ-LNS, and lower VHT monetary incentives. The worst-case scenario simultaneously models an 18% mortality reduction, a 10% increase in the price of SQ-LNS, and higher VHT monetary incentives.

## Discussion

In this study, we developed a framework for modeling the cost and cost-effectiveness of providing SQ-LNS to young children in rural Uganda, a framework that can be adapted and used to make similar sets of estimates in other countries. We estimated that the economic cost of providing SQ-LNS to all children for 12 months, from 6 to 18 months of age, in rural Uganda would be ∼$52 per child (2020 US dollars). Cost-effectiveness estimates were $15,914 per death averted and $242 per DALY averted (undiscounted) or $413 per DALY averted with costs and effects discounted at 3%. Although there is no consensus on a specific cost-effectiveness threshold, the World Health Organization has suggested that interventions with a cost per DALY averted less than gross domestic product (GDP) per capita may be considered very cost-effective (24). Per capita GDP was $822 in Uganda in 2020 (25), so SQ-LNS would be classified as very cost effective. It is important to note, however, that this classification of cost-effectiveness does not imply affordability or policy feasibility.

These results should be interpreted in the context of study strengths and limitations. One strength was the ability to model costs based on cost estimates adapted from a comprehensive costing study of MNP in Uganda. This sets our study apart from many other efforts to model the cost and cost-effectiveness of nutrition interventions that rely on generic unit cost estimates. In addition, our estimates of effects were based on direct evidence of the impact of SQ-LNS on each outcome from recent meta-analyses that included 14-18 randomized controlled trials (mostly in Sub-Saharan Africa) that included a total of >37,000 children. About half of these trials were conducted within existing community-based or clinic-based programs, so the results are relevant to real-world settings. Finally, we used the CHEERS 2022 checklist to ensure we met each relevant criterion for high-quality reporting of our methods and results (26). Because Uganda was not one of the sites in which the trials of SQ-LNS were conducted, one study limitation is that we had to assume that the findings of the meta-analyses would apply to Uganda. Another limitation is that we were not able to estimate costs and cost-effectiveness in urban settings due to the lack of information on the cost of delivering a product like SQ-LNS via platforms that are prevalent in urban areas such as routine health clinic services. As a result, there is uncertainty around the extent to which these results would apply to urban settings or to settings in other countries in which a delivery platform like Uganda’s VHT system does not exist.

We are aware of only one other study that estimated the cost of providing SQ-LNS to children. Hiebert, Phelan (27) estimated the cost of including daily SQ-LNS for children 6-23 months of age as part of an integrated package of maternal and child interventions in rural Niger. They found that the incremental cost of adding 18 months of SQ-LNS delivery per child to the standard of care (interventions that were already being delivered) was $44 per child ($29/year) (2019 US dollars), though this cost was based only on the cost of the product plus freight and is therefore not comparable to our total cost estimates.

In addition to the cost study of MNP from which our cost estimates were derived (8), several studies have modeled the cost and cost-effectiveness of providing various nutrition interventions to young children in Uganda, including Pasricha, Gheorghe (28) who estimated the cost-effectiveness of MNP supplementation, and Shekar, Hyder (29) who estimated the cost-effectiveness of providing complementary food. Several key methodological choices complicate the direct comparison of results across studies. For estimating costs, the chosen perspective and type of costing can significantly influence cost estimates. Cost studies conducted from a societal perspective, as is the case for our study, account for the economic value of all resources used in providing and accessing an intervention for *all* stakeholders. Cost studies conducted from a narrower perspective, for example from the perspective of program implementers or the government, are limited to the costs incurred only by those stakeholders. Similarly, cost studies using economic costing are comprehensive in that all inputs used in providing and accessing an intervention are included, even inputs that are not paid for such as volunteer labor or household time to participate in an intervention, in which case the economic cost of the input is the input’s value in its next best use (i.e., the input’s opportunity cost) (21). This can lead to very different estimates of cost than is the case in studies based only on financial costs, which exclude opportunity costs. These and other intervention characteristics that can influence costs estimates are summarized, by study, in Table 4. After adjusting for differences in study characteristics where possible, estimates from other studies ranged from $22 per child to deliver MNP to $72 per child for the provision of complementary food. Our estimate of $52 per child for SQ-LNS falls within this range. There was wide variation in the percent of the total cost per child attributed to the cost of the product itself, ranging from 18% for MNP based on the Schott, Richardson (8) study, to 77% for complementary food. Our model indicated that product costs represent 42% of the cost of delivering SQ-LNS. If non-product costs of nutrition interventions are underestimated, cost-effectiveness estimates may be overly optimistic.

**Table 4.**
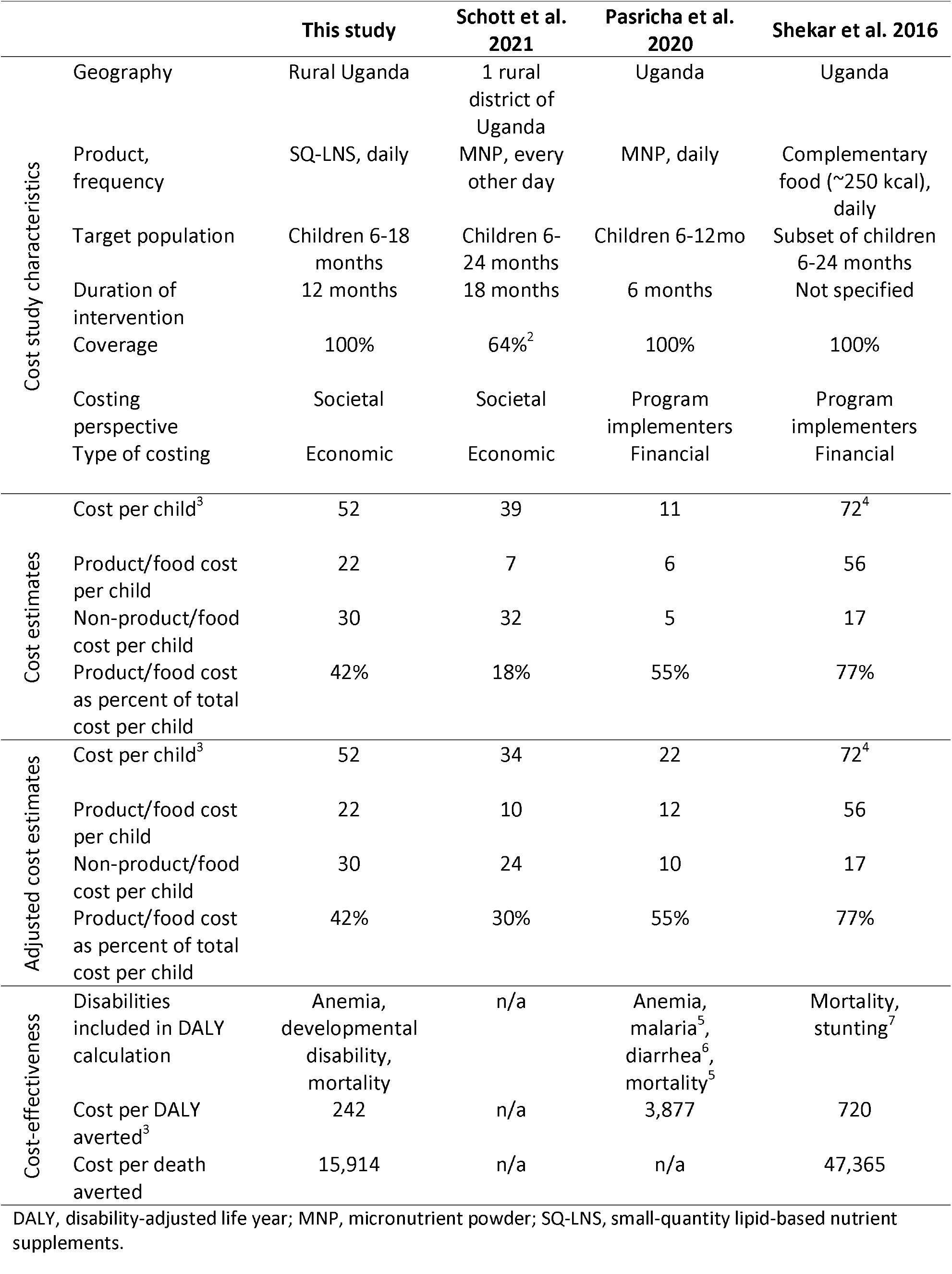
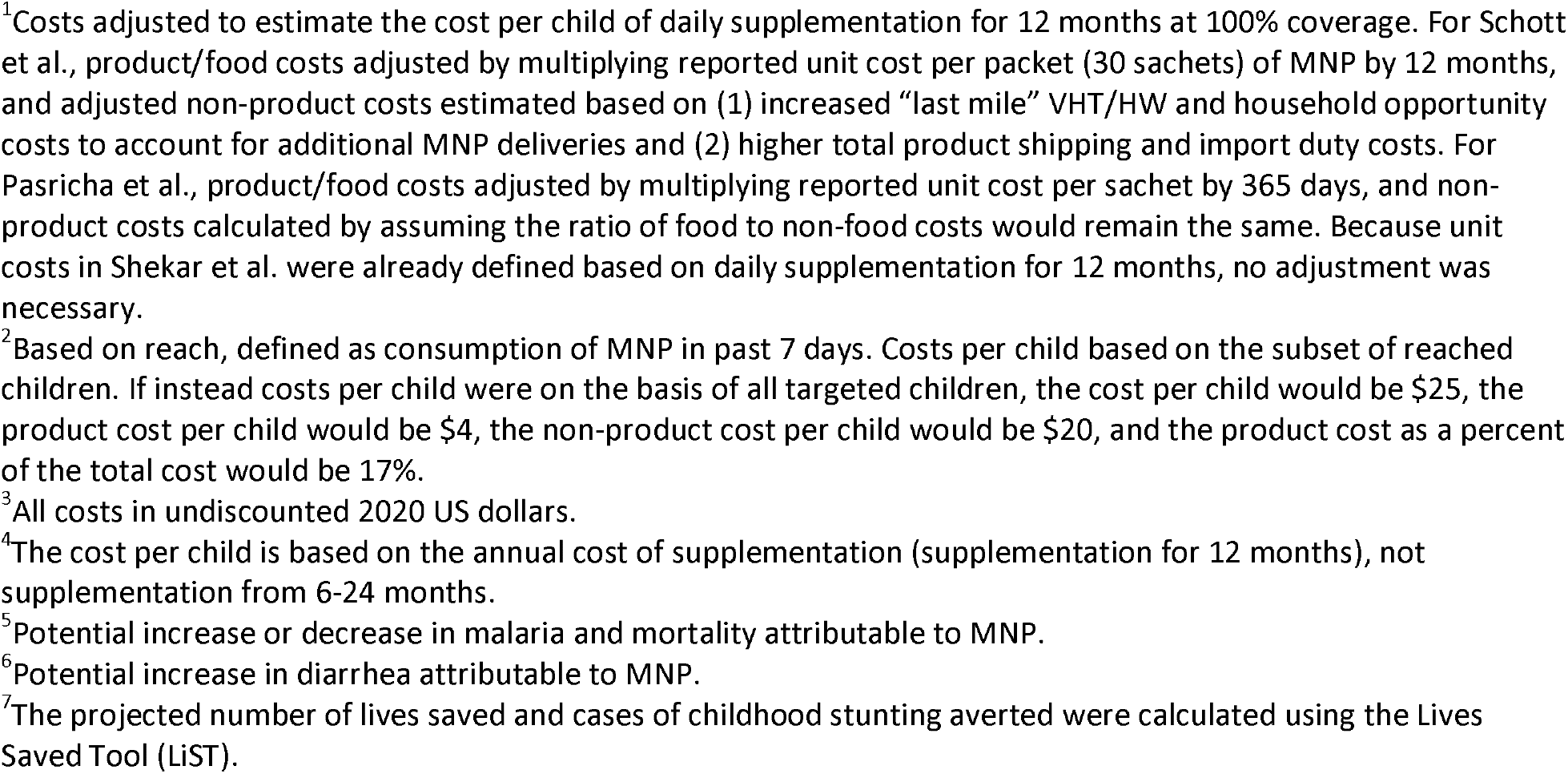
Study-specific costing characteristics and cost, adjusted^1^ cost, and cost-effectiveness estimates

Differences in the methodology used to model the effects of an intervention can also make it difficult to directly compare estimates of cost-effectiveness. The approach we took, which was also taken by Pasricha, Gheorghe (28), was to use direct effects of a specific intervention for all modeling based on systematic reviews of randomized controlled trials of that intervention. This is in contrast to the approach employed by the Investments Framework for Nutrition work (30), including the Shekar, Hyder (29) Uganda study, in which estimated effects of interventions on diseases (morbidity), anemia, wasting and stunting based on systematic reviews of similar interventions were inserted into the Lives Saved Tool (LiST) model to estimate lives saved and other outcomes. The LiST pathways for mortality are defined via cause-specific mortality, which means that the impacts of complementary feeding interventions on child mortality are estimated based on effects on specific diseases, stunting and wasting (31). Our model used direct effects of SQ-LNS on all-cause mortality; we did not have data on cause-specific mortality. Our estimated undiscounted cost per DALY averted was $242 for SQ-LNS, considerably lower than the estimates for MNP ($3,877) and provision of complementary food ($720) in Uganda (Table 4). Our estimated undiscounted cost per death averted was $15,914 for SQ-LNS, much lower than the estimate of $47,365 for provision of complementary food; there is no estimate for MNP because no effect of MNP on mortality has been documented (32). However, it is important to bear in mind that variation in the approaches to cost and effectiveness modeling across these studies could account for some of these differences.

As mentioned, our models assumed that all children would be provided with SQ-LNS for 12 months starting at 6 months of age. We made this assumption because most of the randomized trials in the meta-analyses started SQ-LNS at 6 months and provided the supplements for at least 12 months, so our estimates of effects are mostly relevant to this scenario. In real-world programs, children may begin receiving SQ-LNS later than 6 months, and duration of supplementation might vary. These variables would affect both costs and effectiveness. There is currently insufficient evidence to estimate effectiveness for scenarios in which the age at beginning of supplementation and duration of access to SQ-LNS vary. This is an important gap and is a high priority for further research.

In real-world programs, adherence to the recommended daily consumption of SQ-LNS can also be highly variable. The trials in the SQ-LNS meta-analysis defined adherence in very different ways, so no overall estimate is available. In the program-based trials, the lowest estimate was 37% (33), based on caregiver receipt of SQ-LNS in the previous month, and the highest estimate was 97.4% (34), based on the percentage of caregivers reporting “high adherence” (child consumption > 4 days/week). In 3 other program-based trials, average adherence was 47% (19), 73.5% (35), and 97% (36). Thus, our effectiveness estimates do not assume 100% adherence. Moreover, average study-level adherence generally did not modify the effect of SQ-LNS in the meta-analysis (2). We thus believe that our effectiveness estimates are robust with regard to variation in adherence.

Our model was based on a community-based platform in which community health workers (CHWs) would deliver SQ-LNS every 3 months. More frequent delivery (e.g. every month) would increase costs somewhat, probably similar to the cost increment we modeled for providing a higher VHT incentive in the sensitivity analyses. There is wide variability across and within countries in the performance of CHW/VHT systems, which will influence cost-effectiveness. Some programs may need strengthening to adequately deliver SQ-LNS. On the other hand, the addition of a supplement to be provided by CHWs to caregivers may enhance CHW performance. For example, in Bangladesh (34), CHWs were more likely to conduct the intended monthly home visits when they had a supplement to deliver. Additional implementation science research to evaluate the programmatic impact of adding delivery of supplements such as SQ-LNS to the usual activities of CHWs would be useful.

There are other potential platforms for distribution of SQ-LNS that warrant cost-effectiveness modeling, including provision via health clinics. The integration of SQ-LNS provision into existing programs for wasting prevention and treatment has been evaluated in a health system platform in Burkina Faso (33) and a CHW platform in Mali (19). In both sites, provision of SQ-LNS was a powerful incentive for caregivers to attend monthly screenings for acute malnutrition. If there are cost savings associated with reduced need for treatment for moderate or severe acute malnutrition and for hospitalization, this can be factored into economic analyses of SQ-LNS. Further work is also needed to evaluate the impact of providing SQ-LNS as an incentive to attend health clinics and/or SBCC sessions, with regard to potentially increased uptake of non-nutrition services such as immunizations.

Although our results suggest that provision of SQ-LNS is highly cost-effective, the total cost for a program aimed at all age-eligible children in a given country is high. This is inherent to a food-based intervention that is designed for prevention (rather than treatment) of malnutrition, such as blanket provision of complementary foods. Cost-sharing arrangements in which households or other stakeholders have been called upon to pay some of the cost of goods and services provided to children are not uncommon. Previous work to estimate willingness-to-pay (WTP) for SQ-LNS from several countries in Sub-Saharan Africa suggests that, across countries and methods of eliciting WTP (including hypothetical [money did not change hands] and experimental [money changed hands]), average WTP for SQ-LNS is above the current product price of $0.06 per sachet (37-39). If some rural Ugandan households were willing to cover the cost of SQ-LNS, financing needs for those children would drop to ∼$30 per child, although shifting costs from implementing agencies to households does not change the total societal cost of the intervention.

Although financing needs could be greatly reduced if households covered some or all of the cost of the product, SQ-LNS cost-sharing strategies would need to account for the fact that, in many settings, most households cannot afford to pay the full price of the product. For example, evidence from a market trial in Burkina Faso suggests that, in that context, household persistent demand for SQ-LNS was very limited (40). In addition, expecting households to cover the cost of the product could substantially limit uptake and/or consistent use over time, which has been observed in other settings for preventative health products (41) and would likely have unquantified negative implications for the effectiveness of SQ-LNS on our outcomes of interest. Developing a cost-sharing distribution strategy in which households with the ability to pay are expected to pay while poorer households are provided SQ-LNS free of charge through a voucher system, for example, might be a possibility. This would require research into the cost and feasibility of mechanisms for identifying households who could and could not pay, complemented by research on demand for SQ-LNS, uptake and consistency of use, and benefits under a targeted voucher system. This would provide an empirical basis upon which the cost-effectiveness of such a strategy could be modeled.

Another option for reducing costs would be to target the intervention to the most vulnerable communities, such as those with high levels of stunting, wasting, child mortality or food insecurity. For instance, in Uganda, SQ-LNS could be targeted to rural districts in sub-regions with the highest rates of child mortality. According to the 2016 DHS, of the 15 sub-regions represented in the survey, the worst-off sub-regions in terms of child mortality were West Nile, Busoga, Tooro, Ankole, and Karamoja (42). By matching rural districts to these five sub-regions, we estimated that, if targeting were based on child mortality rates, approximately 35% of children 6-18 months of age in rural Uganda would receive SQ-LNS (396,808 annual average children compared to 1,118,340 children under the untargeted scenario) and while the cost per child would still be ∼$52, the annual average cost of the program would drop by ∼65% (from ∼$58.7 million to ∼$20.8 million per year). In addition to reducing costs, targeting SQ-LNS to children in the most vulnerable communities could enhance effectiveness for certain outcomes. In the SQ-LNS meta-analyses, the impacts of SQ-LNS on development and iron status were greater among children with a higher burden of undernutrition or lower socio-economic status (4, 6).

## Conclusion

Despite increased focus on early childhood malnutrition over the past decade, millions of children in LMICs remain vulnerable to undernutrition (43-45). Strategies that include provision of supplements such as SQ-LNS have been shown to be effective in helping reduce this burden. However, making informed policy decisions aimed at developing a portfolio of effective, cost-effective, and financially feasible interventions requires that robust estimates of intervention program costs and cost-effectiveness be available to policymakers and program planners alongside evidence of effectiveness.

Investing in SQ-LNS has the potential to cost-effectively save child lives and to avert cases of anemia, developmental disability, stunting, and wasting, but the total cost of programs that include SQ-LNS can be substantial. Financing strategies may require innovative financing mechanisms based on stakeholders’ recognition that such programs will enhance child welfare and make fundamental contributions to reducing poverty and promoting overall economic development over the long term. This approach has been used for other types of investments, such as early childhood education (ECE) programs, which contribute to an array of development objectives (46) but are expensive to establish and the maintain ; indeed, multiple stakeholders are generally called upon to fund ECE programs (47). For their part, households recognize these benefits, some of which accrue to them, and are willing to pay for them (48), but requiring resource-poor households to do so can raise equity issues. Similarly, for programs that include SQ-LNS, households may be willing to cover some of the costs (37), though more research is needed on household demand persistence (40). If policymakers choose to invest in SQ-LNS, sustainably financing its incorporation into portfolios of child nutrition interventions may require targeting and/or other strategies to reduce and spread costs among multiple stakeholder groups.

## Data Availability

All data produced in the present study are available upon reasonable request to the authors

## Acknowledgments

The authors gratefully acknowledge Robert Ackatia-Armah for his guidance on the development of the cost model and for providing important Uganda-specific context. We also thank Belinda Richardson and Emily Baker for guiding us through the Uganda micronutrient powder costing study data. We also gratefully acknowledge the coordinating support of Caroline Joyce.

