## Supplemental materials for "The cost-effectiveness of small-quantity lipid-based nutrient supplements for prevention of child death and malnutrition and promotion of healthy development: modeling results for Uganda"

Supplemental Table 1. Base costs and extrapolation dimensions used to adapt Namutumba costs to all other rural districts in Uganda

|  | **Base cost^1^**  **(2020 US dollars)** | **Extrapolation dimension^2^** |
| --- | --- | --- |
| **Programmatic start-up costs** |  |  |
| **Social and behavior change communication** |  |  |
| Personnel | 13,592 | VHT Pop |
| Transport | 3,356 | Space |
| Materials | 58,874 | VHT Pop |
| In-kind incentives | 1,342 | VHT Pop |
| **Capacity building** |  |  |
| Personnel | 6,567 | VHT Pop |
| Transport | 2,190 | Space |
| Materials | 10,759 | VHT Pop |
| In-kind incentives | 10,709 | VHT Pop |
| **Opportunity costs** |  |  |
| **Household opportunity cost "Last Mile"** |  |  |
| Opportunity cost | 0.98 | Child Pop |
| **Recurring programmatic costs** |  |  |
| **Social and behavior change communication** |  |  |
| Personnel | 13,593 | VHT Pop |
| Transport | 3,357 | Space |
| Materials | - | VHT Pop |
| In-kind incentives | 1,343 | VHT Pop |
| **Logistics** |  |  |
| Personnel | 32,035 | VHT Pop |
| Transport | 1,928 | Space |
| Materials | - | Space |
| In-kind incentives | - | Space |
| **Capacity building** |  |  |
| Personnel | 53,715 | VHT Pop |
| Transport | 10,458 | Space |
| Materials | 2,832 | VHT Pop |
| In-kind incentives | 8,998 | VHT Pop |
| **Operational M&E** |  |  |
| Personnel | 15,505 | VHT Pop |
| Transport | 2,676 | Space |
| Materials | 1,251 | VHT Pop |
| In-kind incentives | - | VHT Pop |
| **Overhead and capital costs** |  |  |
| Equipment / Vehicles | 997 | VHT-Child Den |
| Buildings/Structures (desks, etc.) | 21 | VHT Pop |
| Other Materials (office supplies, etc.) | 24 | VHT Pop |
| Overhead | 164 | VHT Pop |

M&E, monitoring and evaluation.

^1^Base costs based on community arm of Schott, Richardson (1) costing study of micronutrient powder in Namutumba district.

^2^VHT pop, the number of village health team (VHT) community health workers per district; Space, the area of each district; Child pop, the number of eligible children per district; VHT-Child Den, the ratio of VHTs to eligible children per district.

Supplemental Table 2. Rural Uganda child population projections

| **Age band** | **2022** | **2023** | **2024** | **2025** | **2026** | **2027** | **2028** | **2029** | **2030** | **2031** |
| --- | --- | --- | --- | --- | --- | --- | --- | --- | --- | --- |
| 6 – 9 mo | 274,758 | 275,647 | 276,574 | 277,656 | 278,915 | 280,257 | 281,314 | 282,283 | 283,246 | 285,200 |
| 9 – 18 mo | 824,274 | 826,942 | 829,722 | 832,968 | 836,744 | 840,770 | 843,942 | 846,850 | 849,738 | 855,601 |
| 18 – 24 mo | 549,516 | 551,295 | 553,148 | 555,312 | 557,829 | 560,513 | 562,628 | 564,567 | 566,492 | 570,400 |

Rural population projections for children 0-59 months based on population projections for children 0-59 months from the Lives Saved Tool (<https://list.spectrumweb.org/>), weighted by projections of the percentage of the Ugandan population classified as rural from the UN World Urbanization Prospects (<https://population.un.org/wup/Download/>). Age band-specific population estimates calculated by assuming a uniform child population size among children 0-59 months.

Supplemental Table 3. Estimated cost of providing daily SQ-LNS to all children in rural Uganda^1^

|  | **2021** | **2022** | **2023** | **2024** | **2025** | **2026** | **2027** | **2028** | **2029** | **2030** | **2031** |
| --- | --- | --- | --- | --- | --- | --- | --- | --- | --- | --- | --- |
| Millions of 2020 US dollars | $10.97 | $56.74 | $56.90 | $57.07 | $57.27 | $57.49 | $57.72 | $57.91 | $58.09 | $58.27 | $58.60 |

^1^Cost estimates based on providing SQ-LNS to all children from 6-12 months of age. Estimates include one year of start-up (2021) in which costs are incurred but benefits do not yet accrue.

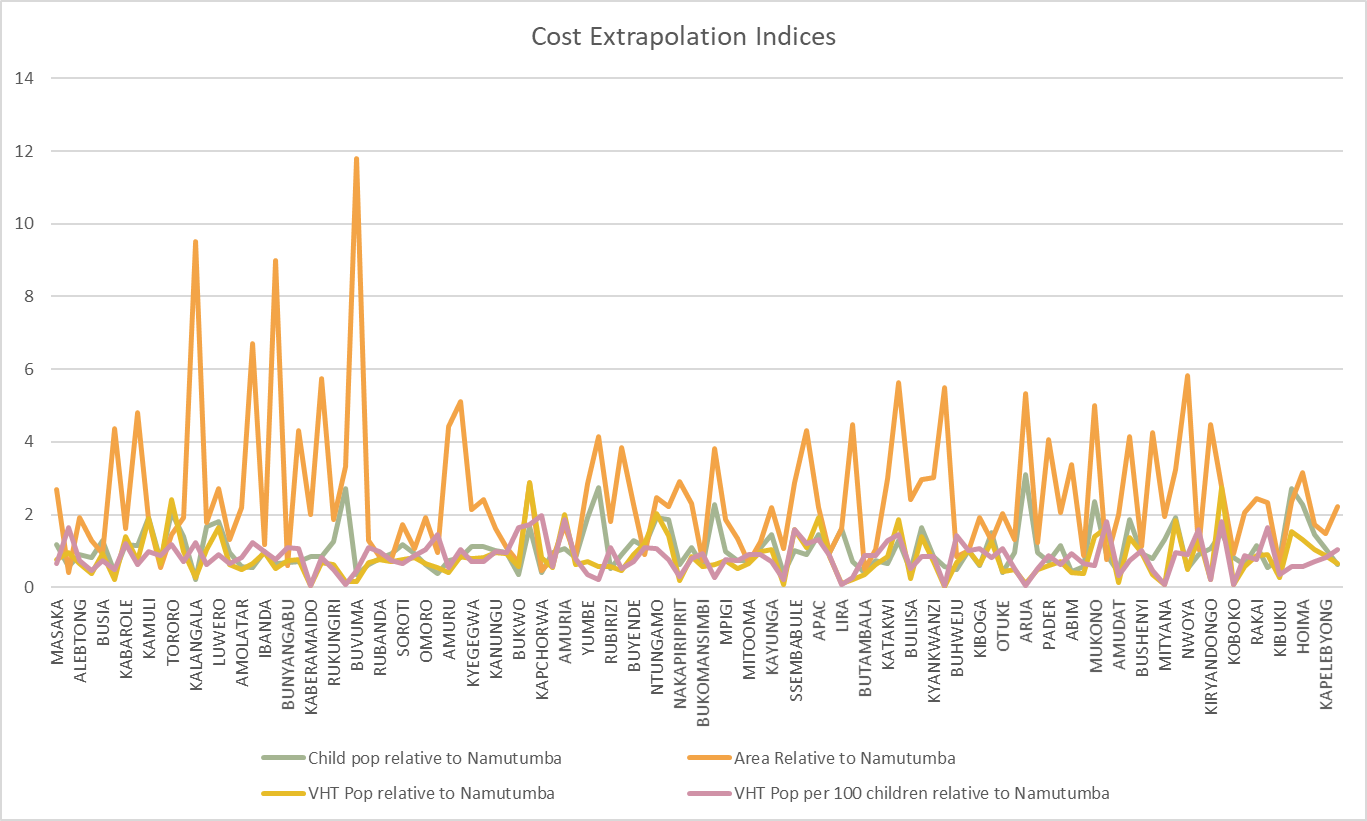

Supplemental Figure 1. Extrapolation indices used to estimate district-level costs based on Namutumba unit costs. Note that the name of all rural districts included in the modeling are not shown on the x-axis.
